# Ultrarare missense and frameshift variants in the *TECTA* gene may involve tectorial membrane in familial Meniere disease

**DOI:** 10.1101/2022.02.18.22270926

**Authors:** Pablo Román-Naranjo, Alberto M. Parra-Perez, Alba Escalera-Balsera, Andrés Soto-Varela, Alvaro Gallego-Martinez, Ismael Aran, Nicolas Perez-Fernandez, David Bächinger, Andreas H. Eckhard, Rocio Gonzalez-Aguado, Lidia Frejo, Jose A. Lopez-Escamez

**Author notes:** Correspondence; (Ext 160).

## Abstract

**Background:** Meniere’s disease (MD) is an inner ear disease defined by episodes of vertigo associated with sensorineural hearing loss initially affecting low- to medium frequencies, tinnitus, and aural fullness. Familial aggregation has been reported in 9-10% of MD patients showing, mostly, an autosomal dominant inheritance pattern with incomplete penetrance. However, familial MD is a genetically heterogeneous disorder and other inheritance patterns have been recently proposed, such as recessive and digenic inheritance involving rare variants in *OTOG* and *MYO7A* genes, respectively. In this study, a familial MD cohort was recruited to identify new candidate genes.

**Methods:** Exome sequencing was performed in 99 individuals (from 77 families) diagnosed with MD according to the diagnostic criteria defined by the Barany Society. Candidate variants were classified based on the ACMG/AMP guidelines, and their effects were evaluated by protein modeling. Standard audiometric evaluations were retrieved, and a case report was made of each family to assess the genotype-phenotype correlations.

**Results:** The *TECTA* gene, which encodes α-tectorin, was highlighted as a candidate for four multicase MD families carrying rare missense heterozygous variants and a short deletion in this gene. Variants in α-tectorin were also found in two additional families with one MD patient and relatives with partial syndromes carrying a missense heterozygous variant and a short deletion. According to the predicted protein model, these variants could affect the stability of α-tectorin.

**Conclusions:** Several MD families were identified carrying rare variants and deletions in the *TECTA* gene, which encodes one of the main proteins of the tectorial membrane (TM). The TM is an extracellular matrix localized over the sensory epithelium mediating the mechanical stimulation of cochlear hair cells, a critical role in the process of hearing. Modifications on the TM stability and the micromechanics involved in the sound-evoked motion of stereocilia might be involved in familial MD.

## BACKGROUND

The tectorial membrane (TM) is an extracellular matrix with a unique structure^1,2^ containing three different collagens and three non-collagenous glycoproteins expressed at high levels just in the inner ear: α-tectorin, β-tectorin, and otogelin^3–6^. Several roles have been suggested for the TM in hearing such as contributing to control of hearing sensitivity by influencing the ionic environment around the hair cell stereocilia^7^. Sound stimulates movement of the hair cells relative to the TM, deflects the stereocilia, and leads to oscillations in the hair cell membrane potential, transducing sound into electrical signals.

Alpha-tectorin (encoded by *TECTA)* is one of the main non-collagenous proteins of the TM, an extracellular matrix that lies over the stereocilia of the hair cells and it is critical for the deflection of the stereocilia and gating of mechanotransduction channels^8^. Mutations in this gene, located in 11q22-q24, are known to cause two phenotypes of non-syndromic autosomal hereditary hearing loss^9^: dominant (Deafness, Autosomal Dominant 8/12; DFNA8/12)^10^ and recessive (Deafness autosomal recessive 21; <u>DFNB21</u>)^11^. To date, over 122 pathogenic or likely pathogenic variants have been identified in the *TECTA* gene that may cause non-syndromic hearing loss (NSHL), not being associated with any other symptoms (http://deafnessvariationdatabase.org/; Release: 4 Jan 2021; Accessed: 29 Oct 2021)^12^.

DFNA8/12, first described in 1998 by Kirschhofer et al.^8^, showed an Austrian family in which 11 individuals covering 4 generations had autosomal dominant NSHL. It was defined by sensorineural hearing loss (SNHL) affecting a broad spectrum of frequencies depending on the domain in which the mutation occurs^13^ with a typical U-shaped audiogram. Its onset could be prelingual or in childhood and the progression of hearing loss can be stable or progressive. *TECTA* mutations are reported in up to 4% of all cases of autosomal dominant NSHL^14^.

Mustapha et al. (1999), described a Lebanese family belonging to the Shiite community in which 9 members presented with a prelingual severe-to-profound sensorineural isolated form of deafness^11^. DFNB21 is defined by a severe-to-profound prelingual hearing loss (70-110dB on all frequencies, most pronounced in mid-frequencies) with a cookie-bite or U-shape audiometric configuration^15^. Nonsense mutations that cause DFNB21 create premature stop codons that truncate the α-tectorin protein, leading to a non-functional version of the protein. A total loss of α-tectorin function alters the structure of the TM in such a way that sound cannot be converted to nerve impulses^16^.

Meniere disease (MD [OMIM 156000]) is an inner ear disorder with an estimated prevalence of 75/100,000 in the Southern European population^17^. It is defined by episodes of vertigo associated with SNHL, initially affecting low-to medium frequencies, tinnitus, and aural fullness^18^. Familial aggregation in MD has been described in 9-10% of European descendant population^19^. Most families show an autosomal dominant inheritance pattern and rare missense variants in several genes have been described in unrelated MD families^20,21^. Nevertheless, none of these variants has been reported in other MD families, supporting genetic heterogeneity in familial MD. Recently in two different studies, enrichment of rare missense variants in the *OTOG* gene was found in 15 unrelated MD families^22^ and, in an exome sequencing study in 62 MD families, other 9 families showed rare heterozygous variants in the *MYO7A* gene associated with the MD phenotype^23^.

Here, we have performed bioinformatic analyses in exome sequencing data obtained from 99 individuals (77 families with MD) and found 7 patients in 4 unrelated families with rare missense variants and a short deletion in the *TECTA* gene. Variants in this gene were also found in two additional families with one MD patient and relatives with partial syndromes carrying a missense heterozygous variant and a short deletion. We suggest that these deletions and missense variants in the *TECTA* gene could change the TM micromechanics involved in the sound-evoked motion of stereocilia in familial MD.

## MATERIALS & METHODS

### Ethics

This study protocol was approved by the local ethics committees (MS/2014/02, Institutional Review Board for Clinical Research, Universidad de Granada, Spain; KEK-ZH-Nr. 2019-01006, Kantonale Ethikkommission, Zurich, Switzerland) and a written informed consent to donate biological samples was obtained from all subjects. The study was carried out according to the principles of the Declaration of Helsinki revised in 2013 for investigation with human subjects.

### Patient assessment, selection, and clinical phenotyping

MD patients were diagnosed and recruited from different Spanish hospitals within the Meniere’s Disease Consortium (MeDiC). Patients were diagnosed according to the diagnostic criteria defined by the International Classification Committee for Vestibular Disorders of the Barany Society in 2015^18^. A complete audiological and vestibular assessment was performed, including magnetic resonance imaging in all cases, to rule out other vestibular diseases that could explain the phenotype. Pure-tone audiograms were retrieved to assess hearing loss since the onset of the disease. A total of 99 Spanish patients with familial MD over 18 years old from 77 different families were selected for exome sequencing.

### DNA extraction and exome sequencing

Blood or saliva samples were collected to obtain DNA as previously described^24^. DNA samples were extracted with QIAamp DNA Mini Kit (Qiagen, Venlo, The Netherlands) and prepIT-L2P (DNA Genotek, Ottawa, Canada), respectively, following the manufacturer’s protocols. DNA concentration and quality parameters were checked by Nanodrop (Thermofisher) and Qubit (Invitrogen) to assess that the samples reach the quality and concentration required for exome sequencing. Additionally, DNA integrity was verified by electrophoresis in a 2% agarose gel.

Exome sequencing was chosen for this study. All exomes were sequenced as previously described^23^. Exome libraries and coding regions were selected using the Agilent SureSelect XT v6 Exome kit (Agilent Technologies, Santa Clara, CA, USA). Sequencing was performed in a NovaSeq 6000 platform (Illumina) with a mean coverage of 100X.

### Processing and dataset generation

All sequenced samples were aligned using the GRCh38/Hg38 reference genome with the maximal exact matches algorithm from Burrows-Wheeler Aligner. For variant calling, we followed standard recommended criteria from GATK. Exome reference alignment, base quality score recalibration, variant calling, and quality filtering pipeline was addressed using Sarek Nextflow pipeline (NF-ACore)^25^. Post-alignment processing was used to remove duplicated reads and the quality of the alignment itself was assessed^26^, genetic variants were then called using the Haplotypecaller function from GATK. After the calling, we merged all the files to generate the MD variant dataset. We performed a variant quality filtering step using the variant quality score recalibration approach recommended by GATK. As a result, two variant call format files were generated, retrieving single nucleotide variants (SNVs) and short insertion and deletions from the sequenced exomes.

### Annotation and prioritization strategy

To annotate the MD variant dataset, we used the Variant Effect Predictor (VEP, Ensembl). Then, we selected minor allele frequency (MAF) thresholds of 0.005 and 0.0005 to identify both autosomal-recessive and autosomal-dominant rare variants in the familial MD cohort, respectively, based on the data from a multi-ethnic study that assessed the pathogenicity of reported NSHL variants^27^. Allelic frequencies were retrieved for the non-Finish European (NFE) population from the Exome Aggregation Consortium (ExAC; N=32,299) and the Genome Aggregation Database (gnomAD; N=33,365) databases, and the Spanish population from the Collaborative Spanish Variant Server (CSVS; N=1,942) database^28^. Candidate variants were then classified according to the American College of Medical Genetics (ACMG) and the Association for Molecular Pathology (AMP) guidelines^29^, according to the specific guidelines for variant interpretation in genetic hearing loss^30^. Additionally, *in-silico* tools such as Sorting Intolerant From Tolerant (SIFT; SIFT<0.05), Polymorphism Phenotyping (Polyphen; Polyphen>0.446), Genomic Evolutionary Rate Profiling (GERP; GERP>2) or Combined Annotated Dependent Depletion (CADD; CADD>15) were used to prioritize and classify each variant according to its predicted pathogenicity.

Lastly, as an additional variant prioritization method, candidate genes carrying rare variants were associated with mammalian phenotypes using the Mouse Genome Database (http://www.informatics.jax.org). Similarly, the Human Phenotype Ontology Project (https://hpo.jax.org/app/) and the Online Mendelian Inheritance in Man (OMIM; https://omim.org/) databases were used to determine associations in humans between candidate genes and phenotypes.

### Statistical analysis

European and Spanish databases such as ExAC, gnomAD, and CSVS were used as references to compare the observed MAF in familial MD. Odds ratios with 95% confidence interval were calculated for each single or set of variants. One-side p-values were corrected for multiple testing following the Bonferroni approach. A corrected p-value < 0.05 was considered statistically significant. The same approach was followed to assess if a combination of variants showed a significant association.

### Candidate variant validation and representation

Regions with prioritized variants were visually inspected using the Integrative Genome Viewer software. Novel variants were validated using Sanger sequencing. Primers used for PCR (Supplementary Table 1) were designed neighboring the regions flanking the variants using the Primer3 v4.1 (http://bioinfo.ut.ee/primer3/), Primer-Blast (https://www.ncbi.nlm.nih.gov/tools/primer-blast/), and the Oligoanalyzer tool (https://eu.idtdna.com/calc/analyzer/). Candidate genes and variants were represented using Illustrator for Biological Sequences Version 1.0^31^.

### Hearing assessment and analysis

To analyze the time course of the hearing profile in familial MD cases with candidate variants, standard audiometric evaluations for air and bone conduction prompted by pure tones from 125 to 8,000Hz were retrieved from their clinical records.

### Alpha-tectorin protein model

The effect of candidate variants on the α-tectorin protein structure was evaluated by protein modeling. The human α-tectorin mature amino acid sequence (from 23 to 2,091) was retrieved from Uniprot entry O75443, containing 10 functional domains. Structural models of the domains were generated using MODELLER (homology modeling)^32^, Robetta-ab^33^ (ab-initio modeling), and AlphaFold2^34^ (ab-initio modeling) methods, and the best ones were assembled using the DEMO method^35^. Each of the generated models and the final assembly were validated using the structure validation algorithms MolProbity^36^, Verify3D^37^, ERRAT^38^, ProSA-Web^39^, and QMEANDisCo^40^. The *in-silico* model was used to predict the stability change (ΔΔG) of the α-tectorin produced by the candidate variants. Hence, we used DynaMut2^41^, MAESTROweb^42^, mCSM^43^, PremPS^44^ and CUPSAT^45^ tools. Variants were classified as neutral when −0.5 < ΔΔG < 0.5^46^. Finally, a prediction of Ca^2+^ ion binding sites was also performed to observe the possible effects of the variants on Ca^2+^ uptake with the metal ion-binding site prediction (MIB) method^47^.

### Variant data and protein structural model submission

All candidate variants in the *TECTA* gene have been submitted to the Clinvar database (http://www.ncbi.nlm.nih.gov/clinvar/). The accession numbers for these variants are as follow: SUB10993075, SUB10994469 and SUB10994413.

The α-tectorin structural model was submitted to the ModelArchive database (https://modelarchive.org/doi/10.5452/ma-xd6ic; Public access after publication. Temporary access code: mwnV7cphoi)

## RESULTS

### Rare missense variants and deletions in the TECTA gene in MD

A total of 34,833 and 20,684 nonsynonymous (missense, start-loss, stop-gain, and stop-loss) or splice-site variants were considered after using, respectively, MAF thresholds of 0.005 and 0.0005 in the familial MD cohort. These rare variants were found in 9,666 genes using the most restrictive MAF threshold and in 12,366 genes using the 0.005 MAF threshold.

After assessing and analyzing those genes with non-singleton variants (i.e., observed in more than one patient), and beyond the genes related to familial MD already published^22,23^, the *TECTA* gene was highlighted as the candidate for 4 multicase MD families carrying rare missense heterozygous variants (F1 – F3) and a short deletion (F4) in this gene. Variants in this gene were also found in two additional families with one MD patient and relatives with partial syndromes carrying a missense heterozygous variant (F5) and a short deletion (F6) (Table 1).

**Table 1:** Rare variants in the *TECTA* gene found in the studied familial MD cohort (N=99).

| Location | Protein change | Info | Exon | Domain | MD family | gnomAD<sub>NFE</sub> MAF | gnomAD<sub>MAX</sub> MAF | ACMG |
| --- | --- | --- | --- | --- | --- | --- | --- | --- |
| 11:121158016T>C | p.Val1494Ala | Missense | 14 | VWFD 4 | F1 & F2 | 8.8x10<sup>-5</sup> | 9.7x10<sup>-5</sup> (AFR) | VUS |
| 11:121152980G>C | p.Cys1402Ser | Missense | 13 | TIL | F3 | 1.5x10<sup>-5</sup> | 4.8x10<sup>-4</sup> (OTH) | VUS |
| 11:121157956AC>A | p.Asn1474LysfsTer91 | Deletion | 14 | - | F4 | Novel | Novel | LP |
| 11:121165368C>T | p.Pro1790Ser | Missense | 17 | - | F5 | Novel | Novel | VUS |
| 11:121189864GC>C | p.Gly2118ProfsTer22 | Deletion | 23 | - | F6 | 0 | 2.4x10<sup>-5</sup> (AFR) | LP |

Reference sequence: NM\_005422.2 (*TECTA*); MAF: Minor allele frequency; NFE: Non-Finish European; MAX: Highest MAF value in gnomAD populations; ACMG: American College of Medical Genetics and Genomics; LP: Likely pathogenic; VUS: Variant of unknown significance

The three variants in all multicase MD families were found clustered in the zonadhesin-like region, between the third trypsin inhibitor-like cysteine-rich (TIL) domain and the fourth von Willebrand factor type D (VWFD) domain. On the other hand, the two variants observed in the two families with partial syndromes were found close to the zona pellucida (Figure 1).

**Figure 1:**
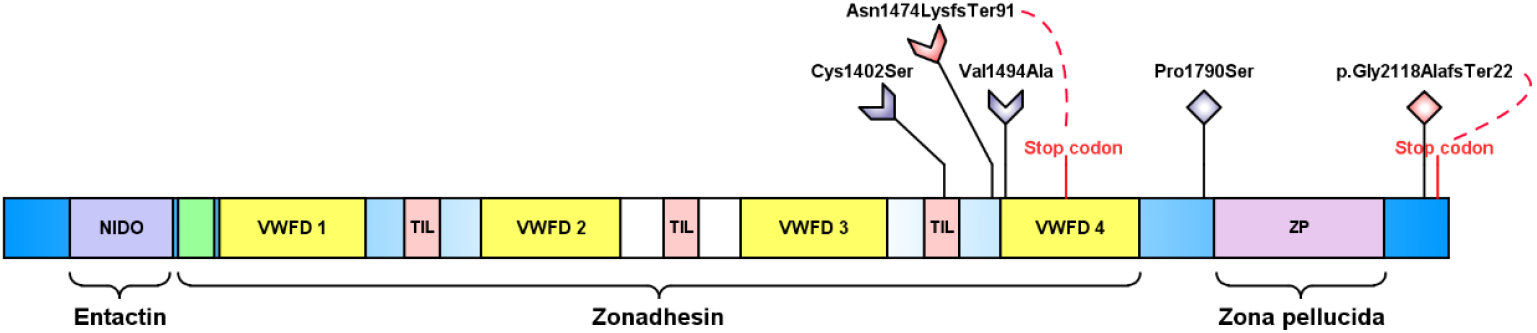
Variants distribution across the α-tectorin domains. The α-tectorin protein has several domains: the NIDO domain, the von Willebrand factor type C (VWFC: colored in green) and type D (VWFD) domains, the trypsin Inhibitor like cysteine rich (TIL) domain and the zona-pellucida (ZP) domains. Three rare missense variants (colored in blue) and two short deletions (colored in red) were found in four multicase MD families (arrow-shaped) and two additional families with one MD patient and relatives with partial syndromes (diamond-shaped). The position of stop codons generated by frameshift short deletions is indicated in red.

Of note, the variant chr11:121158016T>C (p.Val1494Ala) was found in F1, F2, and a sporadic case. The segregation of this variant was confirmed in F1, where it segregated in all three affected MD individuals and the father of III-7, who only suffered from hearing loss. The frequency of this variant in the NFE population from gnomAD is 8.8×10^−5^ and no individuals have been reported as homozygous for the alternate allele. This variant is predicted to impact the protein by several *in-silico* tools (GERP=3.09, PolyPhen=0.954, CADD=24.3) and it was classified as a variant of unknown significance (VUS). Interestingly, a rare variant in chr10:112298172A>C (p.Lys259Gln) in the *TECTB* was found in F2 along with the p.Val1494Ala *TECTA* variant. Moreover, a novel frameshift deletion in chr11:121157956AC>A (p.Asn1474LysfsTer91) found in F4 was classified as likely pathogenic. This deletion resulted in a premature stop codon after 91 amino acids that generates a truncated protein of 1594 residues instead of 2155. The missense variant chr11:121152980G>C (p.Cys1402Ser) found in F3, whose frequency in the NFE population is 1.5×10^−5^, was also predicted to impact the protein (SIFT=0, GERP=5.33, PolyPhen=0.993; CADD=28), and it was classified as VUS according to the ACMG/AMP guidelines.

The two additional variants observed in the families with partial syndromes were classified as VUS. The variant chr11:121165368C>T (p.Pro1790Ser) was found in F5 segregating in two patients, the father with MD and his daughter with only vestibular symptoms. This is a novel variant not observed in gnomAD. Finally, the short deletion chr11:121189864GC>C (p.Gly2118ProfsTer22) observed in F6 results in a premature stop codon at position 2139, generating a slightly shorter protein. This variant has been observed in only one individual (African/African American population) from gnomAD.

### Clinical description of families carrying variants in the TECTA gene

A briefcase report was made of each family to assess the genotype-phenotype correlations. Figures 2 and 3 show the pedigrees and the pure-tone audiograms, respectively, of the six families included in this study. Supplementary Table 2 shows a summary of the clinical information of familial MD patients carrying variants in the *TECTA* gene.

**Figure 2:**
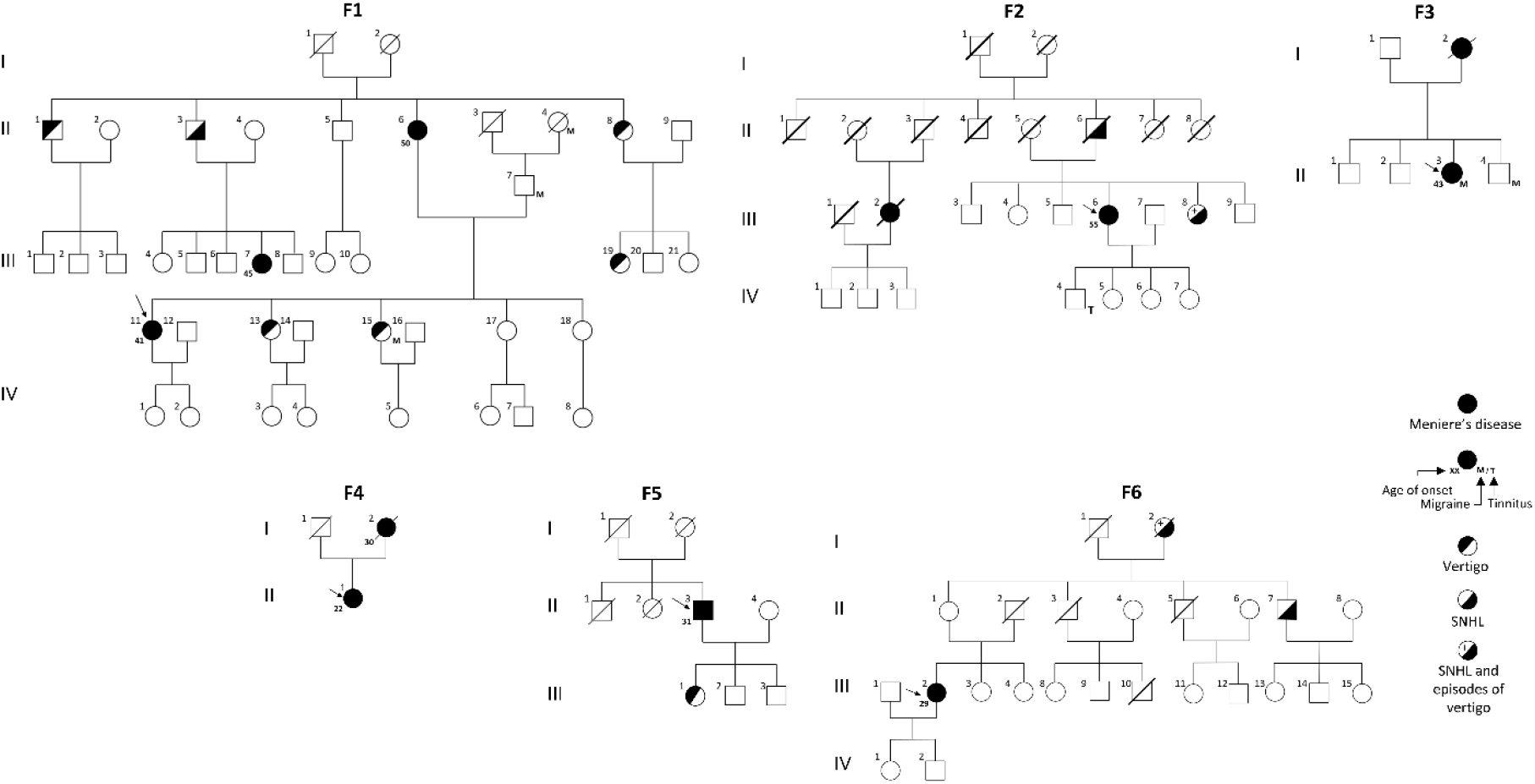
Serial pure tone audiograms for MD patients from the six families included in this study carrying variants in the *TECTA* gene. The different lines in the audiograms show the hearing loss progression across the years (yo). Years after onset are indicated in brackets for each individual.

**Figure 3:**
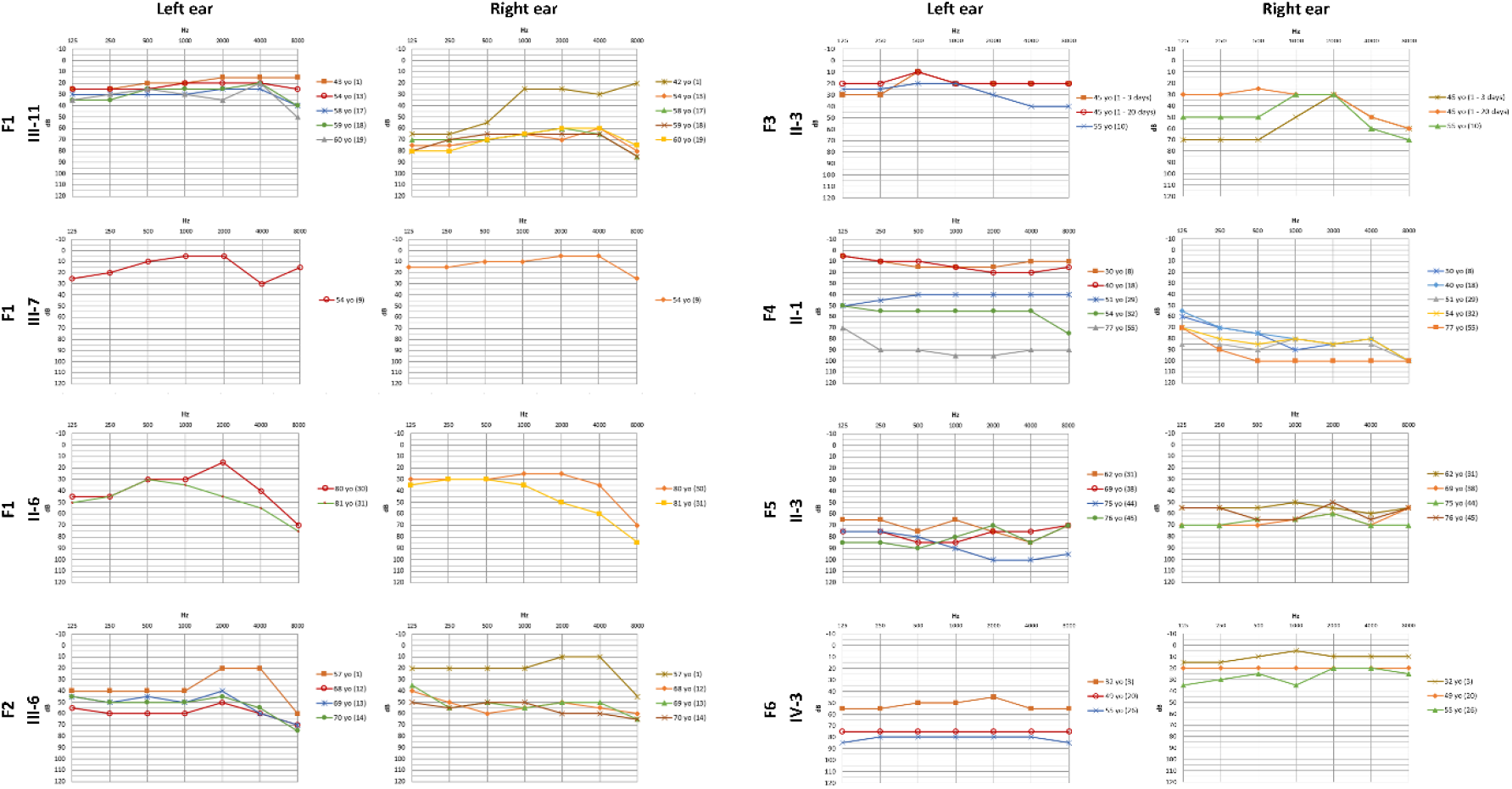
Pedigrees of the six families carrying rare variants and short deletions in the *TECTA* gene.

### Family 1 (F1)

A family consisting of three women in two generations with the complete phenotype was selected for exome sequencing. In this family, six relatives had incomplete phenotypes: five relatives (four women and one man) with recurrent vertigo and one man with SNHL. Additionally, three relatives (two women and one man) also suffered from migraine.

The index case (III-11) was a woman suffering from right SNHL with episodes of vertigo since her 40s. However, in few years, the hearing loss evolved to bilateral SNHL. Her parent (II-6), who is currently in her 80s, was diagnosed with left-sided MD when she was 46-50 years old. The last time she was examined the patient presented bilateral hearing loss with a normal gain in the video head impulse test (vHIT). Finally, the relative III-7 was diagnosed with bilateral MD. Currently, III-7 is 51-55 years old and, when she was evaluated, she did not show hearing nor vestibular symptoms, showing normal hearing and vestibulo-ocular reflexes.

### Family 2 (F2)

This family consists of two female cousins with MD in one generation (III-2 and III-6) where only III-6 (index patient) was available for exome sequencing. III-6 is in her 70s and suffers from unilateral (left ear) hearing loss since she was 51-55. The disease progressed from unilateral to bilateral MD when she was 61-65 years old, suffering from that moment mainly vestibular symptoms. Some of her relatives had partial syndromes: her parent (II-6) suffered from tinnitus, a trait that is also present in her child (IV-4). In addition, one of her relatives (II-8) presented SNHL and vertigo episodes who did not meet the diagnostic criteria for MD.

### Family 3 (F3)

The third family consists of two relatives with MD, the parent (I-2) and her child (II-3), only being available for exome sequencing II-3. There are no intermediate phenotypes in this family, neither hearing loss, nor vertigo or tinnitus. Two relatives of II-3 have a history of high blood pressure. Additionally, one of them was diagnosed with cardiac valvulopathy and the other with psoriasis. The index case (II-3) is a woman currently in her 50s suffering from fluctuating hearing loss and tinnitus in her right ear since she was 41-46. She experienced her first vertigo episode associated with fluctuating hearing loss at 45 years old. During the next 5 months, she suffered from 8 episodes of vertigo, 4 of them lasting more than two hours. Right canal paresis (62%) was evidenced by bithermal caloric testing. The patient started treatment with acetazolamide (250-500 mg daily) without any improvement in her symptoms. Next, two intratympanic instillations of dexamethasone (23mg/ml) were administrated in the right ear, with an improvement in control of vertigo. In a 10-year-follow-up, this patient suffered from 4 episodes of vertigo. Her parent (I-2) suffered from left-sided MD, but it progressed from unilateral to bilateral MD with severe hearing loss at the end of the disease. This patient used a combination of trimetazidine and vestibular sedatives to control vertigo attacks.

### Family 4 (F4)

A fourth family consisting of two relatives with MD in two generations (parent and child) was studied. Only the child (II-1, index patient) was available for exome sequencing. Her parent, who died at the age of 36-40, suffered from unilateral MD since her thirties.

II-1 is currently a woman in her 80s suffering from bilateral MD. At the age of 21-25, the disease started as unilateral, affecting her right ear. We could not assess the hearing loss progression in this ear since the first audiogram we could recover was when the patient already suffered from severe SNHL affecting all frequencies. Over the years, the disease progressed affecting both ears, estimating the onset of the left hearing loss between the ages of 40 and 50. Hearing loss progressed in both ears showing a flat-type audiogram in her 70s. The vestibular assessment was performed at 76-80 year old by vHIT testing, showing a bilateral vestibular hypofunction. Due to a tympanostomy tube, caloric testing could not be performed. In this same year, the patient got a cochlear implant in her right ear.

### Family 5 (F5)

A family consisting of a man with MD (II-3; index patient) and his child (III-1) with partial syndromes. II-3 is a man in his 70s suffering from bilateral MD since he was 31-35. We could not evaluate the hearing progression on this patient since the first audiogram we were able to retrieve was when he was 61-65 and the disease had progressed for more than 30 years. At that time, hearing loss was already affecting all frequencies in both ears. His child, currently in her 50s, reported episodes of vertigo accompanied by tinnitus and headache during the last 5 years. However, she did not show a hearing loss. None of the other relatives reported MD nor partial syndromes: II-1 and III-2 suffered from high blood pressure and II-2 suffered from diabetes mellitus type 2.

### Family 6 (F6)

The last family consisted of a woman with MD (IV-3) and two other relatives with partial syndromes (a woman (I-1) and a man (II-7)). The index patient (IV-3) is a woman in her 50s suffering from recurrent vertigo associated with tinnitus and aural fullness (left ear) since she was 26-30. This patient suffers from profound unilateral hearing loss affecting all frequencies since the first years of the disease. According to the familial pedigree, these traits (i.e., hearing loss or vertigo) were exclusively identified on the maternal side of the proband: her relative I-2 suffered from unknown etiology hearing loss and vertigo and her relative II-7 only suffered from hearing loss.

### Sporadic patient

We have also identified a patient (woman; 71-75 years old) with sporadic MD carrying the same missense variant in the *TECTA* gene as F1 and F2 patients. This patient reported her first episode of vertigo when she was 61-65, when she had two prolonged episodes of vertigo, referring to a sensation of spinning motion, right-sided hearing loss, and vegetative symptoms. One week after her second vertigo episode, the audiogram showed normal thresholds except for a slight drop in high frequencies.

Few years later, the patient returned to the hospital reporting a new episode of vertigo, beginning with a sensation of spinning motion followed by tinnitus and hearing loss in her right ear. At that time, the audiogram showed a moderate to severe SNHL in her right ear involving all frequencies. During that year, the patient reported new episodes of vertigo. Caloric testing was conducted, showing normal results. Vertigo attacks were controlled with steroids and betahistine.

When she was 71-75, the patient reported multiple episodes of vertigo lasting several hours with worsening hearing loss and aural fullness. The patient followed her treatment of betahistine and a salt-reduced diet was recommended. During the last years, the patient has not reported any vestibular symptoms, although she has referred tinnitus in her right ear.

### Protein modeling

The α-tectorin protein model was obtained by modeling the protein domains using the AlphaFold2 method and assembled with the DEMO method (Figure 4). According to the geometrical validation results (Supplementary Table 3), we have obtained a reliable model compared to structures solved by experimental techniques at the geometric level. This model was used to predict the impact of SNVs on protein stability and Ca^2+^ binding sites.

**Figure 4:**
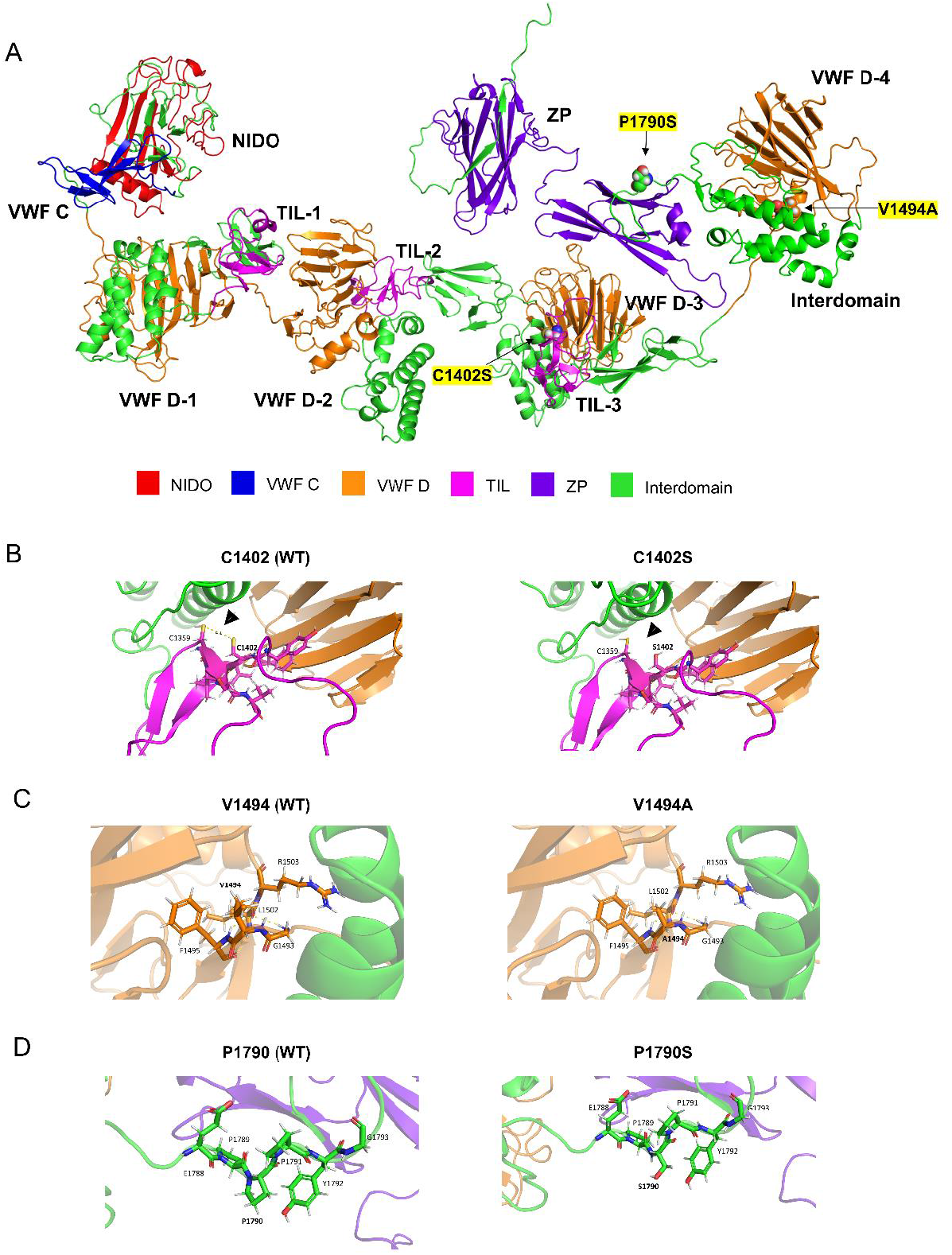
Predicted effect of variants on the α-tectorin structure. A) Mature α-tectorin model showing the different domains that form the protein and the positions of the missense variants found in this study (indicated with black arrows). B) TIL-3 domain of WT α-tectorin (left) and C1402S mutant (right). A disulfide bridge between C1359 and C1402 is predicted in the WT protein (indicated with an arrow). This disulfide bridge disappears in the mutated protein, where appears a weak polar interaction between C1359 and S1402. C) VWF D-4 domain of WT α-tectorin (left) and V1494A mutant (right). D) Interdomain upstream the ZP domain of α-tectorin WT (left) and P1790S mutant (right). Yellow dashed lines represent polar interactions between residues within 4 Å from the mutated residue. Arrows show new or missing bonds. VWF D: Von Willebrand factor type D; ZP: Zona Pellucida.

p.Cys1402Ser (Cys1380Ser, in the mature protein without the signal peptide and the final propeptide), p.Val1494Ala (Val1472Ala), and p.Pro1790Ser (Pro1768Ser) variants were predicted *in-silico* to change the overall stability of the α-tectorin (Supplementary Table 5). Consistent with the used methods, variants p.Cys1402Ser found in F3 and p.Val1494Ala found in F1 and F2 were classified as destabilizing variants. In contrast, most of the used predictors classified the novel variant p.Pro1790Ser found in F5 as a neutral variant according to the predicted perturbation on protein stability. Four probable Ca^2+^ binding sites were predicted in the α-tectorin VWFD domain where the p.Pro1790Ser is located, however, this variant appears to not affect them (Supplementary Table 4 and Supplementary Figure 2).

## DISCUSSION

Our findings support those rare variations in the *TECTA* gene may affect the stability of the protein α-tectorin in patients with familial MD suggesting a novel mechanism involving the TM in MD. The TM is an acellular structure that covers the sensory epithelium of the cochlea in the organ of Corti. This membrane, which is composed of a mixture of collagenous and non-collagenous proteins, plays a critical role in the process of hearing mediating the mechanical stimulation of cochlear hair cells^48^. Thus, variants in these non-collagenous proteins of the TM, namely otogelin (*OTOG*), otogelin-like (*OTOGL*), α-tectorin (*TECTA*), β-tectorin (*TECTB*), and the carcinoembryonic antigen cell adhesion molecule 16 (*CEACAM16*), may result in different types of autosomal dominant or autosomal recessive hearing loss in humans: DFNB18B^49^, DFNB84B^50^, DFNA8/12^10^, DNFB21^11^, DFNA4B^51^ or DFNB113^52^. In this study, we have identified 3 rare missense variants and 2 deletions in the *TECTA* gene in 4 multicase MD families and 2 families with MD and partial syndromes by exome sequencing. Of note, one of these rare variants (p.Val1494Ala) was found in two unrelated families segregating the MD phenotype, and two deletions generating a truncated form of the α-tectorin protein in other two families. These variants in α-tectorin could change the TM stability, disturbing the sound-evoked motion of hair cell stereocilia in familial MD.

Familial aggregation is reported in 9-10% of MD patients showing, mostly, an autosomal dominant inheritance pattern^19^. However, other inheritance patterns have been recently proposed involving rare variants in *OTOG* and *MYO7A* genes. Enrichment of rare variants was found in the *OTOG* gene in 15 families with MD, suggesting a recessive inheritance pattern mediated by compound heterozygous variants in this gene^22^. The *OTOG* gene encodes otogelin, an extracellular protein that participates in the otolith tethering in the otolithic membrane, the TM attachment crowns, and the horizontal top connectors between stereocilia^6,53^. On the other hand, it was suggested a potential digenic inheritance pattern in familial MD mediated by rare missense variants in *MYO7A* and other genes encoding for proteins involved in the organization of the stereocilia links (e.g., *CDH23, PCDH15*, or *ADGRV1*)^23^. The *MYO7A* gene encodes the myosin VIIA protein, a motor protein involved in the formation of ankle links and tips links in the stereocilia of hair cells^54^. These two genes, *OTOG* and *MYO7A*, are closely related to the *TECTA* gene since they are highly expressed in the hair cell stereocilia and involved in the interaction with the TM. Rare variants in any of these genes could reduce the stability of the stereocilia proteins, the attachments of the apical crowns of the stereocilia to the TM or generate structural changes in the TM. This may lead to a sudden onset of spontaneous oscillations in a subset of hair bundles, resulting in a spurious signal that may cause abnormal depolarization of hair cells and fluctuation in hearing in MD.

The *TECTA* gene encodes the α-tectorin protein, one of the major non-collagenous proteins of the TM^48^. This large protein contains 10 functional domains divided into three major regions: 1) an entactin-like (NIDO) region; 2) a larger middle region, the zonadhesin region containing a von Willebrand factor type C (VWFC) domain, four VWFD domains, and three TIL domains; and 3) the zona pellucida region^5^. Although there is little evidence, several molecular models involving α-tectorin, β-tectorin, and CEACAM16 have been proposed for the formation of the striated-sheet matrix, strands of filaments that organize the collagen fibers in the TM^55,56^. Both, α- and β-tectorin contain a ZP region, a polymerization domain that could mediate the formation of either homomeric or heteromeric filaments crosslinked by CEACAM16. Studies in mice lacking functional TECTA (*Tecta*^*ΔENT/ΔENT*^) showed a TM absent of striated-sheet matrix and completely detached from the organ of Corti^16^. Furthermore, several models carrying heterozygous variants in different *Tecta* domains were generated as models from human *TECTA* variants. These variants disrupted the structure of the covernet fibrils, the marginal band, the Hensen’s stripe, and the Kimura’s membrane^57,58^. The structural consequences of variants in the *TECTA* gene showed in this study are expected to be milder compared to the phenotype showed in the *Tecta*^*ΔENT/ΔENT*^ mice model, resembling more the phenotypes exhibited by those mice models carrying heterozygous variants in *Tecta*.

The SNVs described in this study were investigated at the functional level according to the predicted α-tectorin protein model. The variant p.Val1494Ala was found in F1, F2 and a sporadic case of our sporadic MD cohort, being classified as VUS according to the ACMG/AMP criteria. Although this variant was located in the VWFD domain, a domain that can bind Ca^2+^, the variant p.Val1494Ala seems to have no effect in the Ca^2+^ binding sites predicted using the protein model structure. In addition, based on the protein stability predictors, this variant produces a slightly destabilizing effect on α-tectorin. The missense variant p.Cys1402Ser found in F3 was classified as a VUS according to the ACMG/AMP guidelines. The change of cysteine to serine at residue 1402 can lead to the breaking of the Cys1359-Cys1402 disulfide bond. The suggested disulfide bond disruption could destabilize the striated-sheet matrix structure of the TM, causing progressive hearing loss^59^.

In addition, a novel frameshift deletion p.Asn1474LysfsTer91 was found in F4, resulting in the absence of the fourth VWFD domain and the ZP domain. ZP and VWFD domains have been suggested to be involved in the TM matrix assembly through the formation of homomeric filaments of α-tectorin or heteromeric filaments with β-tectorin^5,60^. In addition, the TM has been suggested as a reservoir for Ca^2+^ cations, which are needed to control the mechanotransduction channel in the stereocilia of hair cells^7^. Apparently, behind this function are the VFWD domains of α-tectorin and otogelin proteins, which can bind Ca^2+^ ions^61^. Thus, the short deletion found in F4 could potentially decrease Ca^2+^ uptake by the VWFD domain and prevent the formation of α-tectorin homodimers and α-tectorin/β-tectorin heterodimers.

Rare variants in the *TECTA* gene were also detected in two families with MD and partial syndromes. The novel variant p.Pro1790Ser in F5 was classified as a neutral variant in terms of protein stability perturbation by 4 out of 6 prediction tools. Nevertheless, we cannot discard a deleterious effect of this variant for a reason different from stability. In fact, this variant, which affects the ZA-ZP interdomain of α-tectorin, is only one amino acid residue downstream of the p.Pro1791Arg variant, firstly described by Hildebrand *et al*. in an American family suffering from prelingual mid-frequency SNHL^14^. Later, this variant was also found in a man suffering from progressive, postlingual SNHL^62^. Finally, although the p.Gly2118ProfsTer22 found in F6 occurs in the last exon, which is part of the α-tectorin propeptide, this region contains elements essential for its function. The α-tectorin suffers a post-translational modification in which it is tethered to the membrane via glycosylphosphatidylinositol (GPI), a process that seems to be needed to prevent the diffusion of secreted TM component. The C-terminal, region where the short deletion found in F6 is located, contains the GPI anchorage signal^63^. As a result, this frameshift deletion could be involved in an alteration of the TM during its formation by the modification of the GPI anchorage signal and leading to the clinical phenotype.

The phenotype observed in the six families carrying rare heterozygous variants in the *TECTA* gene was described. In general, we observed a trend towards postlingual hearing loss worsening with age involving all frequencies. Conversely, we did not identify a different audiometric profile according to the region where the variants were localized, nor the cookie-bite/U-shaped audiogram typically observed in patients carrying recessive mutations in *TECTA*^9,15,64^. Interestingly, the patients carrying frameshift deletions in this study (i.e., F4 &F6) presented an earlier age of onset compared to those patients carrying rare missense variants, suggesting a more significant alteration in the TM matrix that could correlate to an earlier occurrence of the phenotype.

Unlike the hearing loss phenotype, the association between variants in the *TECTA* gene and vestibular dysfunction has not been established since most patients carrying variants in α-tectorin do not show a vestibular phenotype. Nevertheless, some of these patients reported episodic vertigo or showed vestibular hyporeflexia^60,65,66^. Furthermore, the expression of α-tectorin in the vestibular system has been demonstrated in several mouse models, being detected in the saccule and the utricule^67^. Mice lacking α-tectorin showed reduced otoconial membranes with no obvious behavioral defects^16^. In addition, a mechanism for otolith tethering requiring otogelin and α-tectorin has been proposed in a zebrafish model^53^. In humans, Calzada *et al*. found that all patients who experienced drop attacks showed disrupted utricular otolithic membranes. Additionally, they showed that the average thickness of the utricular otolithic membrane in patients with MD was 11.45 micrometers, while in normal tissue the mean thickness was 38 micrometers^68^. This reduced thickness in the otolithic membrane could be associated with changes in the structure of the proteins involved in the otolithic membrane, including α-tectorin. Nevertheless, since the role of α-tectorin in the human vestibular system is not completely clear, we cannot discard other genes or epigenetic factors that could modulate the vestibular phenotype in MD patients.

The results achieved in this study seem to be in line with the results obtained in most recent genetic studies about familial MD, where rare variants in *MYO7A* and *OTOG* gene were suggested to modify the stability or the interactions of different proteins in the apical surface of the sensory epithelia, such as hair cells (stereocilia) or the TM^22,23^. In this study, the presence of rare missense variants and frameshift deletions in the *TECTA* gene in six unrelated families with MD suggests a role of this gene in the pathophysiology of the disease. However, because of the lack of a reliable association between α-tectorin and the vestibular function, we consider that additional variants and genes may contribute to the vestibular phenotype in MD.

## Data Availability

The data that support the findings of this study are available from the corresponding author upon reasonable request.

## Acknowledgments

The authors appreciate the participation of all patients with MD and their families in this study. This project was partially funded by H2020-SC1-2019-848261 (UNITI), CECEU PY20-00303 (EPIMEN), European Regional Funds B-CTS-68-UGR20 (M3N-OMIC) and the Schmieder-Bohrisch Foundation (Geneva, Switzerland). DB is supported by a national MD-PhD scholarship from the Swiss National Science Foundation (SNSF). AHE is supported by a career development grant (“Filling the Gap”) from the University of Zurich, Switzerland.

## LIST OF ABBREVIATIONS

ΔΔG: Stability change
ACMG: American College of Medical Genetics
AMP: Association for Molecular Pathology
CADD: Combined Annotated Dependent Depletion
CSVS: Collaborative Spanish Variant Server
ExAC: Exome Aggregation Consortium
GERP: Genomic Evolutionary Rate Profiling
gnomAD: Genome Aggregation Database
GPI: Glycosylphosphatidylinositol
MAF: Minor allele frequency
MD: Meniere’s disease
MeDiC: Meniere’s Disease Consortium
MIB: Metal ion-binding site prediction
NFE: Non-Finish European
NSHL: Non-syndromic hearing loss
Polyphen: Polymorphism Phenotyping
SIFT: Sorting Intolerant From Tolerant
SNHL: Sensorineural hearing loss
SNVs: Single nucleotide variants
TIL: Trypsin Inhibitor like cysteine-rich
TM: Tectorial membrane
VEP: Variant Effect Predictor
vHIT: video head impulse test
VWFC: von Willebrand factor type
C VWFD: von Willebrand factor type
D ZP: Zona-pellucida

## SUPPLEMENTARY MATERIAL

## SUPPLEMENTARY TABLES

**Supplementary Table 1:**
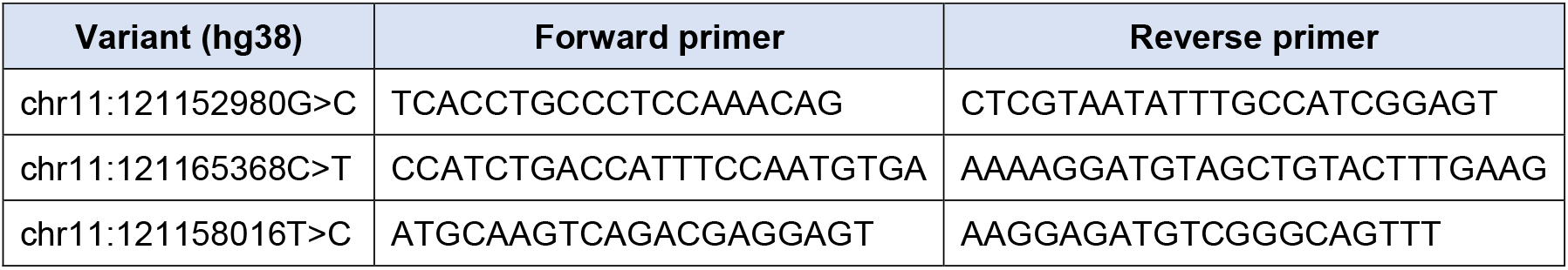
Pair of primers used to validate candidate single nucleotide variants by Sanger Sequencing in the *TECTA* gene.

**Supplementary Table 2:** Summary of the clinical information of familial MD patients carrying variants in the *TECTA* gene.

| Families |  | F1 | F2 | F3 | F4 | F5 | F6 |
| --- | --- | --- | --- | --- | --- | --- | --- |
| Genetic variants | *TECTA* gene | p.Val1494Ala | p.Val1494Ala | p.Cys1402Ser | p.Asn1474LysfsTer91 | p.Pro1790Ser | p.Gly2118ProfsTer22 |
| MD patients | Index | III-11 | III-6 | II-3 | II-1 | II-3 | III-2 |
|  | Other | II-6 III-7 | III-2 | I-2 | I-2 | - | - |
| Relatives with incomplete phenotype | Episodic vertigo | II-1, II-8, III-13, III-15, III-19 | III-8 | - | - | III-1 | I-2 |
|  | Hearing loss | II-3 | II-6, III-8 | - | - | - | I-2, II-7 |
| Clinical data of index patient | Sex | Female | Female | Female | Female | Male | Female |
|  | Laterality (ear) | Unilateral (Right) | Bilateral | Unilateral (Right) | Bilateral | Bilateral | Unilateral (Left) |
|  | Age of onset | 41-45 | 51-55 | 41-45 | 21-25 | 31-25 | 26-30 |
|  | Autoimmune diseases | No | No | No | No | No | No |

**Supplementary Table 3:**
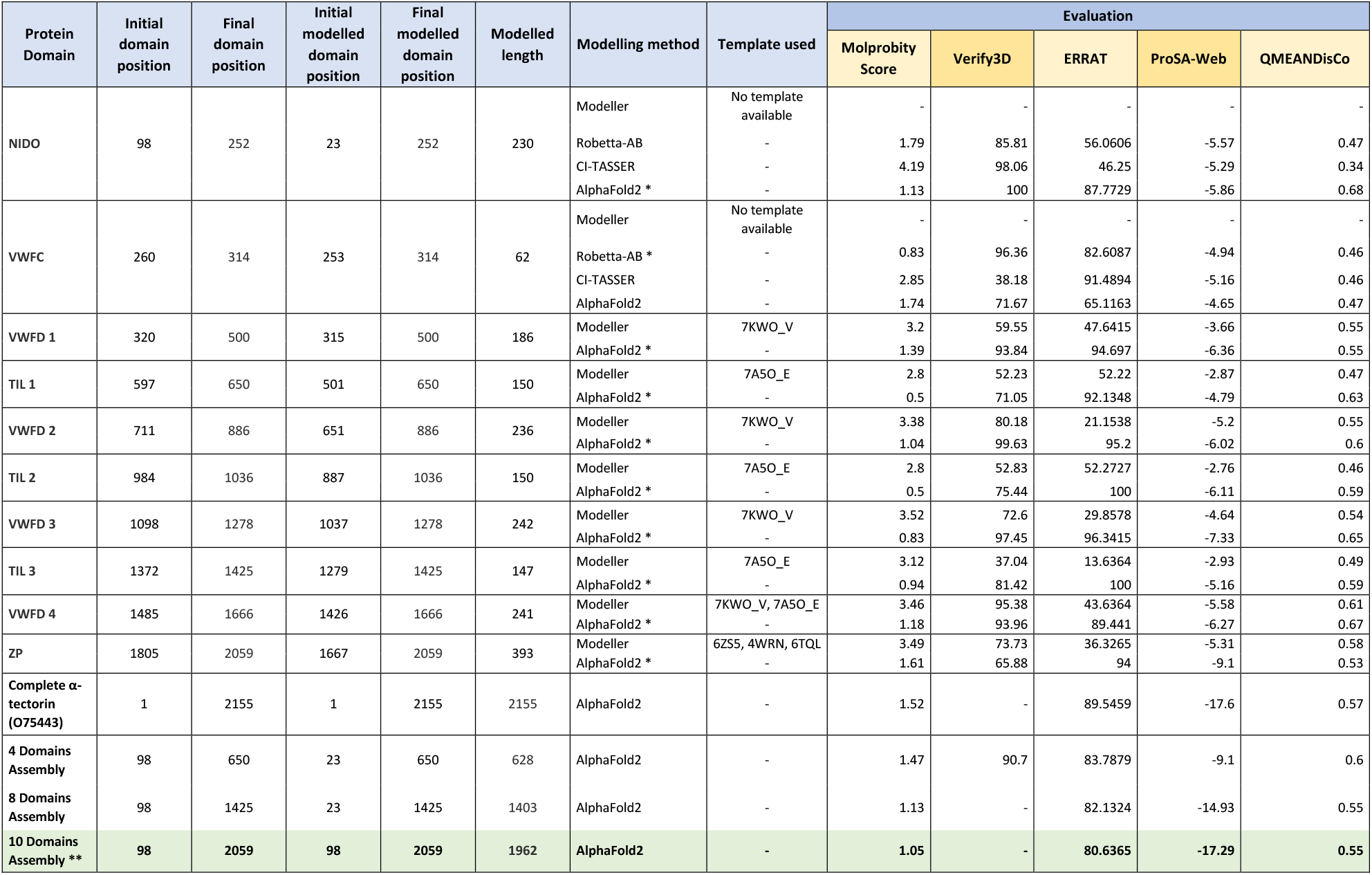
Predicted structural model evaluation of each of the α-tectorin domains, the 4-, 8-, and 10-domain assemblies of the protein using the DEMO method and the α-tectorin structural model predicted by Alphafold2 (located at https://alphafold.ebi.ac.uk/entry/O75443). Molprobity Score, Verify3D, ERRAT, ProSA-web and QMEANDisCo metrics were used in the evaluation. Molprobity Score is a weighted logarithmic combination of different geometric scores such as clashscore, percentage of unfavoured Ramachandran and percentage of bad sidechain rotamers, giving a number that reflects the crystallographic resolution at which those values would be expected. Lower values of Molprobity Score are better. Verify3D determines the compatibility of an atomic model (3D) with its own amino acid sequence (1D) by assigning a structural type based on its location and environment. A higher score indicated high-quality of the structure. The overall quality factor, ERRAT, analyses the statistics of interactions between the different types of atoms and plots the value of the error function calculated by a comparison with highly refined structure statistics. As the generally accepted range for a high-quality model is >50, this analysis revealed that the backbone conformation and nonbonded interactions of all models were within the scope of a high-quality model. In the ProSA-web tool, the score is z-score defined as the energy separation between the native fold and the average of an ensemble of the misfolds in standard deviation units of the database. A z-score outside a range characteristic for native proteins of similar sizes indicated an erroneous structure. In this case, each model is in the range. Finally, QMEANDisCo evaluates the agreement of pairwise distances between residues with sets of distance constraints extracted from structures homologous to the evaluated model, so the higher the score the better the model. * Selected structural model domain to build the assembly and model the whole protein. ** Model used for evaluating the impact of the variants found in this study.

**Supplementary Table 4:** Probable Ca2+ binding sites in the α-tectorin VWFD4 protein structure.

| Binding site | α-tectorin residue number | Amino acid | Score (MIB) |
| --- | --- | --- | --- |
| 1 | 1507 | ALA | 2.096 |
|  | 1508 | ASN | 2.096 |
| 2 | 1530 | ASN | 1.682 |
|  | 1531 | PHE | 1.682 |
|  | 1532 | ASP | 1.682 |
| 3 | 1624 | ASN | 2.143 |
|  | 1626 | ASN | 2.143 |
|  | 1627 | GLY | 2.099 |
|  | 1628 | ASP | 2.143 |
|  | 1630 | THR | 1.641 |
|  | 1631 | ASP | 2.143 |
|  | 1632 | ASP | 2.143 |
|  | 1499 | ASP | 1.746 |
| 4 | 1664 | SER | 2.097 |
|  | 1666 | ASN | 2.097 |

**Supplementary Table 5:** Predicted α-tectorin stability perturbation caused by missense variants found in this study. Global protein stability change prediction (kcal/mol) in the α-tectorin model using different ΔΔGpred prediction methods. For Maestro-web and PremPS, ΔΔGpred < 0.0 indicates a stabilizing mutation. On the other hand, for mCSM, CUPSAT and DynaMut2, ΔΔGpred > 0.0 indicates a stabilizing mutation. Variants were considered neutral in terms of protein stability perturbation when −0.5 < ΔΔGpred < 0.5.

| Predictor | Variants |  |  |
| --- | --- | --- | --- |
|  | p.Cys1402Ser | p.Val1494Ala | p.Pro1790Ser |
| Maestro-web (kcal/mol) | Neutral (0.110) | Neutral (0.056) | Neutral (0.022) |
| mCSM (stability) (kcal/mol) | Destabilizing (-1.254) | Destabilizing (-1.099) | Destabilizing (-0.545) |
| CUPSAT thermal (kcal/mol) | Destabilizing (-13.02) | Destabilizing (-0.57) | Stabilizing (2.23) |
| CUPSAT denaturants (kcal/mol) | Stabilizing (12.46) | Stabilizing (0.79) | Neutral (-0.4) |
| DynaMut2 (kcal/mol) | Destabilizing (-0.54) | Destabilizing (-1.29) | Neutral (-0.25) |
| PremPS (kcal/mol) | Destabilizing (2.2) | Destabilizing (0.6) | Neutral (0.43) |

## SUPPLEMENTARY FIGURES

**Supplementary Figure 1:**
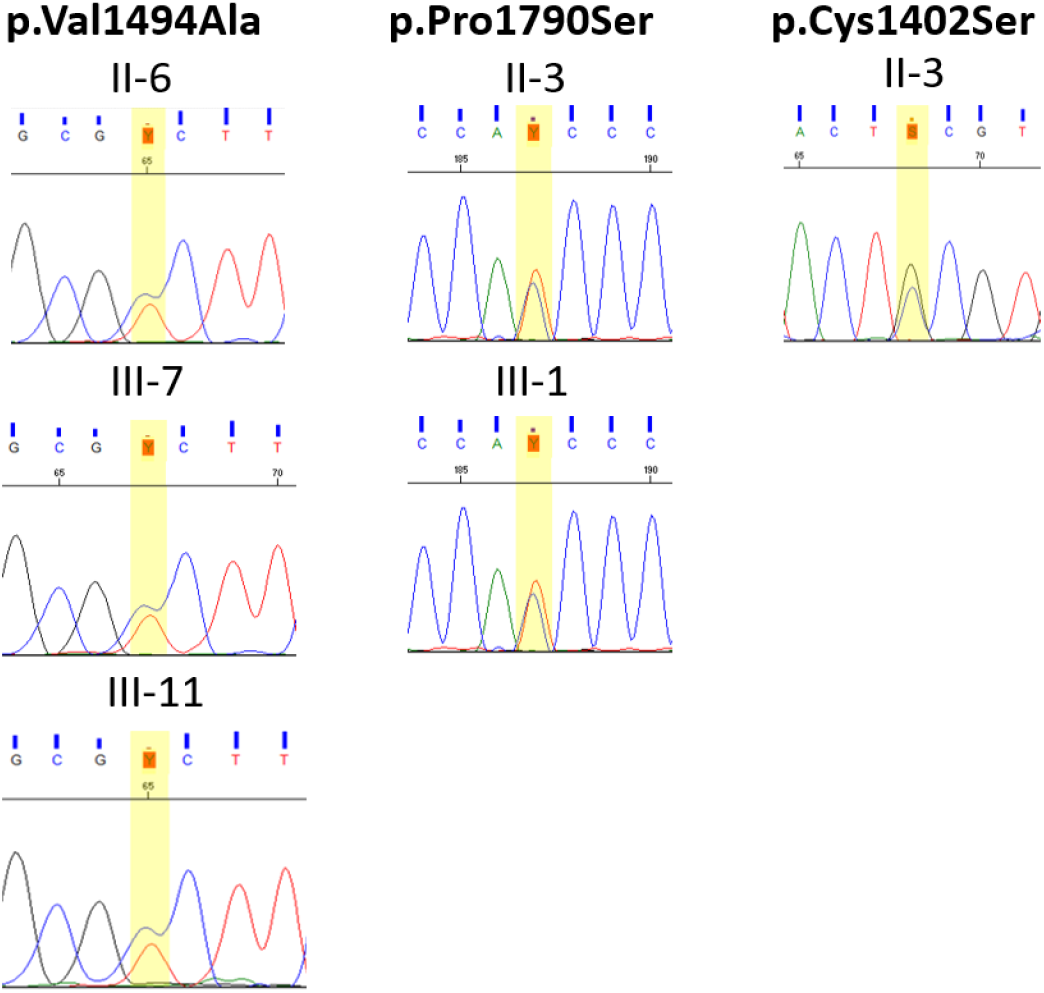
Validation of candidate variants by Sanger sequencing.

**Supplementary Figure 2:**
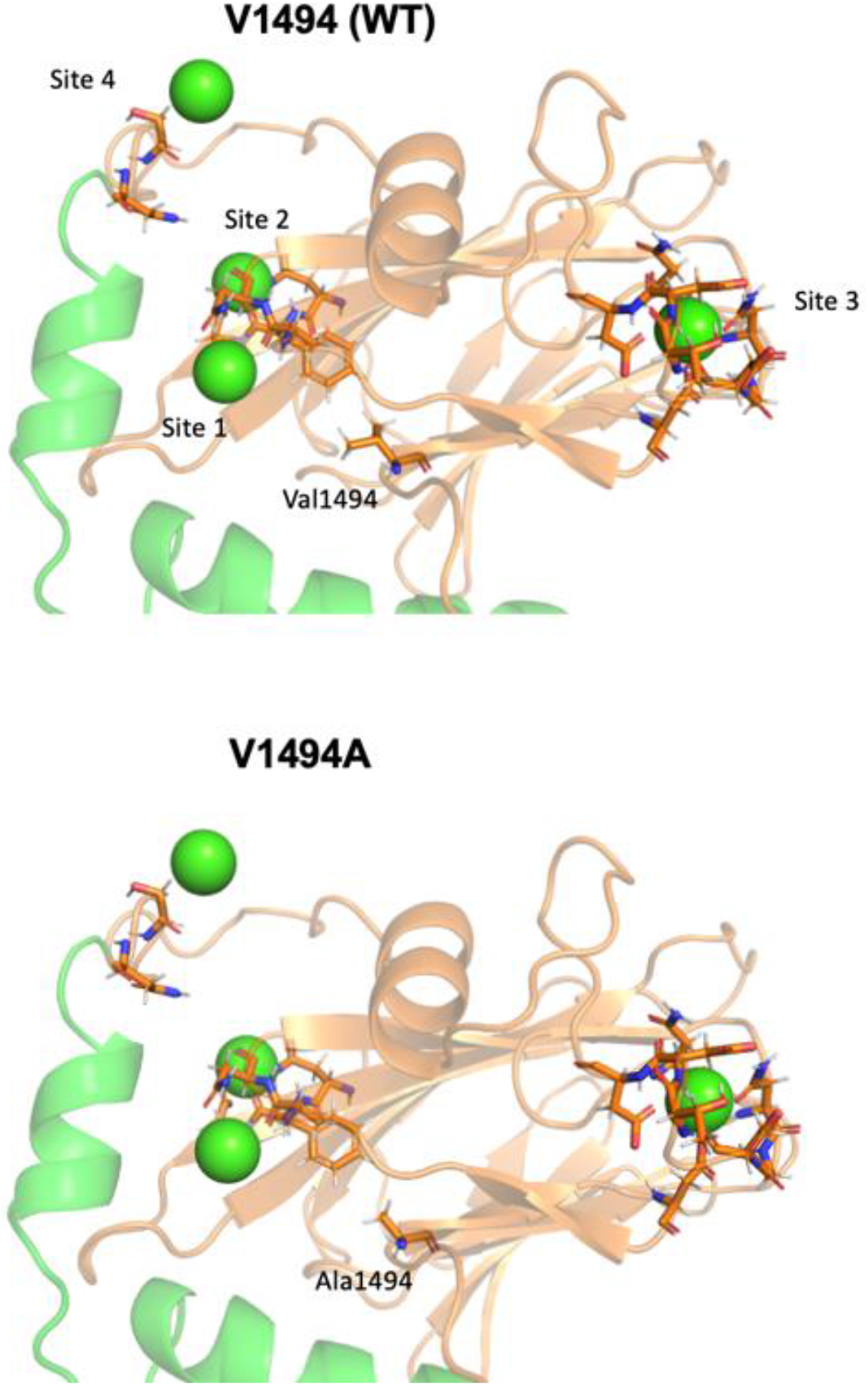
The four predicted Ca2+ binding sites in the α-tectorin VWFD4 protein structure (Supplementary Table 3 – coloured in this figure in green) located near the variant p.Val1494Ala, found in F1, F2 and a sporadic patient with MD.

